# Identification of differences in the magnitude and specificity of SARS-CoV-2 nucleocapsid antibody responses in naturally infected and vaccinated individuals

**DOI:** 10.1101/2023.01.05.23284247

**Authors:** Pradeep D. Pushpakumara, Chandima Jeewandara, Farha Bary, Deshan Madushanka, Lahiru Perera, Inoka Sepali Aberathna, Thashmi Nimasha, Jeewantha Jayamali, Thushali Ranasinghe, Heshan Kuruppu, Saubhagya Danasekara, Ananda Wijewickrama, Graham S. Ogg, Gathsaurie Neelika Malavige

## Abstract

**Background:** As there are limited data on B cell epitopes for the nucleocapsid protein in SARS-CoV-2, we sought to identify the immunodominant regions within the N protein, recognized by patients with varying severity of natural infection with the Wuhan strain (WT), delta, omicron and in those who received the Sinopharm vaccines, which is an inactivated, whole virus vaccine.

**Methods:** Using overlapping peptides representing the N protein, with an in-house ELISA, we mapped the immunodominant regions within the N protein, in seronegative (n=30), WT infected (n=30), delta infected (n=30), omicron infected+vaccinated (n=20) and Sinopharm (BBIBP-CorV) vaccinees (n=30). We then investigated the sensitivity and specificity of these immunodominant regions and analysed their conservation with other SARS-CoV-2 variants of concern, seasonal human coronaviruses and bat Sarbecoviruses. We then investigated the kinetics of responses to these regions in those with varying severity of acute COVID-19.

**Results:** We identified four immunodominant regions aa 29-52, aa 155-178, aa 274 to 297 and aa 365 to 388, were highly conserved within SARS-CoV-2 and the bat coronaviruses. The magnitude of responses to these regions varied based on the infecting SARS-CoV-2 variants, with WT infected individuals predominantly recognizing aa155 to 178 regions, delta infected individuals and vaccinated+omicron infected individuals predominantly recognizing regions aa 29 to 52 and aa 274 to 294 regions. Sinopharm vaccinees recognized all four regions, with the magnitude of responses significantly lower than other groups. >80% of individuals gave responses above the positive cut-off threshold to many of the four regions, with some differences with individuals who were infected with different VoCs. These regions were found to be 100% specific, as none of the seronegative individuals gave any responses.

**Conclusions:** N-protein specific responses appear to be detectable in over 90% of those who were naturally infected or vaccinated with a whole virus inactivated vaccine, with responses mainly directed against four regions of the protein, which were highly conserved. As these regions were highly specific with high sensitivity, they have a potential to be used to develop diagnostic assays and to be used in development of vaccines.

## Introduction

The SARS-CoV-2 virus continues to evolve, giving rise to more immune evasive and more transmissible variants, which continue to drive outbreaks globally [21]. Although variants such as omicron (BA.1) were thought to initially cause milder illness, the sub-lineages that subsequently emerged such as BA.2, were associated with more severe disease in certain populations [30]. In fact, BA.2 outbreaks in the United States and in Hong Kong resulted in several fold higher mortality rates than seen during the delta outbreaks in many countries [13]. Many factors could contribute to the differences in mortality rates and hospitalization rates during different outbreaks in different countries such as co-morbidities, age, vaccination rates of a population, the proportion of individuals naturally infected, COVID-19 control measures, better treatment modalities, infra-structure to manage hospitalized patients and seasonal changes [2,5,18]. Among all the factors that have contributed to a reduction in mortality rates, COVID-19 vaccines, are likely to be one of the single most important factors that were responsible for this reduction [2,29].

While neutralization antibodies (Nabs) have shown to associate with protection against severe disease when infected with the SARS-CoV-2 [1,12], the mRNA COVID-19 vaccines appear to induce higher levels of Nabs compared to other vaccines [15]. However, there is emerging evidence that nucleocapsid (N) protein specific antibody responses may be protective based on data in animal models [8]. Indeed, a high frequency of polyfunctional T cell responses specific for certain epitopes within the N protein was found to associate with milder illness [22] and N protein specific antibody responses were detected earlier in infection and were present at detectable levels in a larger proportion of individuals compared to spike protein specific antibody responses [6].

The N protein is one of the most abundant, highly conserved RNA-binding proteins, which plays an important role in the packing of the SARS-CoV-2 genome [3]. It plays an important role in the regulation of the virus replication cycle, inhibits interferon response and induced apoptosis [3]. The N protein, which spans 419 amino acids, consists of five domains and all five have shown to bind to RNA [6]. The region starting from the 388 amino acid position was found to induce a high frequency of immune responses in patients with acute COVID-19 (from the Wuhan strain) and was found to be 100% specific to detect infection with SARS-CoV-2. Although the N protein is an important T cell and antibody target, the main immunodominant regions within this protein, targeted by antibodies has not been extensively studied. For instance, although the N protein is highly conserved, as it is an important antibody target, certain mutations in SARS-CoV-2 variants of concern (VoC), can give rise to differences in the magnitude of antibody responses to certain regions. Therefore, we sought to identify the immunodominant regions within the N protein, recognized by patients with varying severity of natural infection with the Wuhan strain (WT), delta, omicron and in those who received the Sinopharm vaccines, which is an inactivated, whole virus vaccine.

## Methods

### Participants for identification of immunodominant regions within the N protein

Blood samples from healthy adult volunteers who were either vaccinated or naturally infected with SARS-CoV-2 were obtained following informed written consent. Serum separated from blood samples were used to assess antibody responses to the overlapping N peptides in the following groups of individuals.

A. SARS-CoV-2 seronegative negative individuals (n=30) prior to COVID-19 vaccination (negative)
B. Unvaccinated individuals who were naturally infected (n=30) with the SARS-CoV-2 wild type/Wuhan strain (WT) from day 14 to 21 from day of onset of symptoms. 5/30 of them had severe illness and 25/30 had mild. Clinical disease severity was classified according to the WHO COVID-19 disease severity classification [31]. (WT)
C. Unvaccinated individuals who were naturally infected with the SARS-CoV-2 delta variant (n=30), 7 to 21 days from the onset of symptoms. All individuals had mild infection. (delta)
D. Those who were vaccinated or who possibly had prior infection (infection status unknown) in those who were subsequently infected with omicron (n=20) days 14 to 21 since onset of illness. (Omicron+vaccinated)
E. Sinopharm (BBIBP-CorV) vaccine recipients 2 weeks post second dose (n=30) (Sinopharm)

#### Ethics statement

Blood samples were obtained following informed written consent. Ethics approval was obtained from the Ethics Review Committee of University of Sri Jayewardenepura.

### Participants for assessing the kinetics of antibody responses to immunodominant regions of N protein

After identification of immunodominant regions within the N protein, the responses to these regions were further assessed in the groups of individuals described above. However, smaller numbers were included in the analysis due to limitations in the sample volume available.

A. SARS-CoV-2 seronegative negative individuals (n=15) prior to COVID-19 vaccination
B. Unvaccinated individuals who were naturally infected (n=12) with the SARS-CoV-2 wild type/Wuhan strain (WT) from day 14 to 21 since onset of illness
C. Unvaccinated individuals who were naturally infected with the SARS-CoV-2 delta variant (n=12), with mild illness, from day 7 to 14 since onset of illness.
D. Those who were vaccinated (different vaccines) or who possibly had prior infection (infection status unknown) in those who were infected with omicron (n=22), 14 to 21 days since onset of illness. All participated individuals had mild infection.
E. Sinopharm (BBIBP-CorV) vaccine recipients 2 weeks post second dose (n=12)
F. Uninfected individuals who received COVID-19 vaccines, which only contain the SARS-CoV-2 spike protein, 3 months since obtaining the second dose. AZD1222 (ChAdOx1) (n=10), Moderna (mRNA-1273) (n=10) and Sputnik V (Gam-COVID) (n=10). This was to assess the specificity of the responses to the immunodominant N peptides.

### Participants with acute infection due to SARS-CoV-2 WT virus

Adult patients who were acute infected with the SARS-CoV-2 virus and had mild illness (n=16) or severe illness (n=9) during acute stage (<7 days since onset of symptoms) and during late infection (21 to 28 days since onset of symptoms) were recruited following informed written consent, to compare antibody responses against four immunodominant regions between individuals with mild and severe disease during early and late stages of the illness. Clinical disease severity was classified according to the WHO COVID-19 disease severity classification [31].

### N protein peptide array

Overlapping peptides representing the N protein of SARS-CoV-2 virus (USA-WA1/2020 strain of SARS-CoV-2; QHO60601) was obtained through BEI Resources, NIAID, NIH: Peptide Array, SARS-Related Coronavirus 2 Nucleocapsid (N) Protein, NR-52404. The whole peptide array consists of 59 overlapping peptides, which overlap by 10aa to 17aa and 10aa with the adjacent peptide. All peptides were dissolved in appropriate solvent mentioned by the manufacturer. Initially, all 59 peptides were pooled in to 4 pools. Namely, pool 1 (peptide 1 to 15), pool 2 (peptide 16 to 30), pool 3 (peptide 31-45) and pool 4 (peptide 46-59). Antibody responses to the peptides which gave the highest responses were further assessed.

### Identification of SARS-CoV-2 serostatus of the participants

The Wantai SARS-CoV-2 total antibody ELISA (Beijing Wantai Biological Pharmacy Enterprise, China) was used to identify the presence of antibodies (IgM, IgG and IgA) to the receptor binding domain (RBD) of the virus. The specificity of this assay in the Sri Lankan population was found to be 100% [17] SARS-CoV-2. Those who tested negative for the presence of total antibodies to the RBD by this assay, were considered to be seronegative.

In those who had received the spike protein contained vaccines, the presence of asymptomatic infection with the virus was assessed by the presence of N protein specific antibodies. This was done by using the Elecsys® Anti-SARS-CoV-2 electrochemiluminescence immunoassay (Cat: 09 203 095 190, Roche Diagnostics, Germany) using the Cobas e 411 analyzer (Roche Diagnostics, Germany). A Cutoff index (COI) ≥1.0 was interpreted as reactive and COI <1.00 was considered non-reactive as indicated by the manufacturer.

### Measuring ACE2 blocking antibodies by the surrogate virus neutralizing test (sVNT)

ACE2 blocking antibodies were measured using the sVNT assay which measures the percentage of inhibition of binding of the RBD of the S protein to recombinant ACE2 (Genscript Biotech, USA). Inhibition percentage ≥ 25% in a sample was considered as positive for Nabs in the Sri Lankan population as previously described [26].

### Identification of SARS-CoV-2 variants in individuals who were naturally infected with SARS-CoV-2

In this study we recruited individuals infected with the WT, delta and omicron. All those who were considered to be infected with the WT had a confirmed SARS-CoV-2 infection (PCR positive) between in March to May 2020, when other VoC were not detected. Infection with either delta or omicron was identified by carrying out genomic sequencing using either the Oxford Nanopore (ONT) or the Illumina platforms as previously [23].

### In-house ELISA to determine IgG antibody responses to the SARS-CoV-2 overlapping peptides of the N protein

Ninety-six-well microtitre plates (Thermofisher, USA, Pierce™ Cat: 15031) were coated with the overlapping peptide representing the different pools and incubated overnight at4□°C. The peptides were diluted in bicarbonate/carbonate coating buffer (pH 9.6) and the final concentration of each peptide was 1μg/100μl. The plates were blocked with PBS with 2% (w/v) bovine serum albumin (Sigma Aldrich, Germany, Cat: A7030) and incubated for 2 hours at room temperature and washed before incubation with serum samples diluted 1:500 in 1% BSA. After an incubation of 30□min at room temperature, the plates were washed, and incubated with biotinylated goat anti-human IgG antibody (Mabtech, Sweden, Cat: 3820-4-250) diluted 1:1000 in 1% BSA. After a 30-minute incubation at room temperature, the plates were washed and further incubated with Streptavidin–HRP (Mabtech (Sweden) Cat: 3310-9) diluted 1:1000 in 1% BSA solution for 30 minutes. After washing the plates, the TMB ELISA substrate solution (Mabtech, Sweden, Cat: 3652-F10) was added at 100μl/well and the plates were incubated in the dark for 10□minutes at room temperature. The reaction was stopped by adding 2M H_2_SO_4_ (Sigma Aldrich, Germany, Cat: 339741) and absorbance values were read at 450nm. Optic density (OD) values above the mean± 3SD of the OD values of the sera from SARS-CoV-2 seronegative individuals was considered as a positive response for a particular peptide or a pool of peptides.

### Statistical Analysis

GraphPad Prism version 9 was used for statistical analysis. As the data were not normally distributed, differences in means were compared using the Mann-Whitney U test (two tailed). The descriptive statistics including the mean and frequencies were used to compare antibody responses of individual peptides. Kruskal-Wallis test was used to determine the differences between the antibody levels (indicated by the OD value) in the four peptide pools (pool 1, 2, 3, and 4). and four immunodominant regions (P5/6, P23/24, P40/41, and P53/54). If Kruskal-Wallis test was significant, a post hoc test (Dunn test) was done to identify which group or groups different from others. Spearman’s correlation coefficient was used to determine the correlation between antibody responses against immunodominant regions of N protein and neutralizing antibodies.

## Results

### Identification immunodominant regions within the N protein of SARS-CoV-2

We initially tested the four pools of the SARS CoV-2 overlapping peptides of the N protein in the cohorts A to E. WT, delta, omicron infected and sinopharm vaccinated individual’s antibody responses were significantly different in the four overlapping pools of peptides (Figure 1A to 1D). The number of individuals included in each of the cohorts that tested positive for the different pools is shown in table 1. Omicron infected + vaccinated individuals (who were vaccinated) had the highest positivity rates (>75%) for all four peptide pools.

**Table 1:** The number of individuals in different cohorts who gave a positive response to different overlapping peptide pools of the N protein.

| SARS-CoV-2 Variant | Number of individuals who gave a positive antibody response |  |  |  |  |
| --- | --- | --- | --- | --- | --- |
|  | Pool 1 | Pool 2 | Pool 3 | Pool 4 | At least one pool |
| WT (n=30) | 9 (30%) | 15 (50%) | 2 (6.7%) | 3 (10%) | 18/30<br>(60%) |
| Delta (n=30) | 10 (33.3%) | 9 (30%) | 17 (56.7%) | 16 (53.3%) | 22/30<br>(73.3%) |
| Omicron + vaccinated<br>(n=20) | 16 (80%) | 15 (75%) | 18 (90%) | 16 (80%) | 18/20<br>(90%) |
| Sinopharm (n=30) | 11 (36.7%) | 7 (23.3%) | 5 (16.7%) | 4 (13.3%) | 14/30<br>(46.7%) |

**Figure 1:**
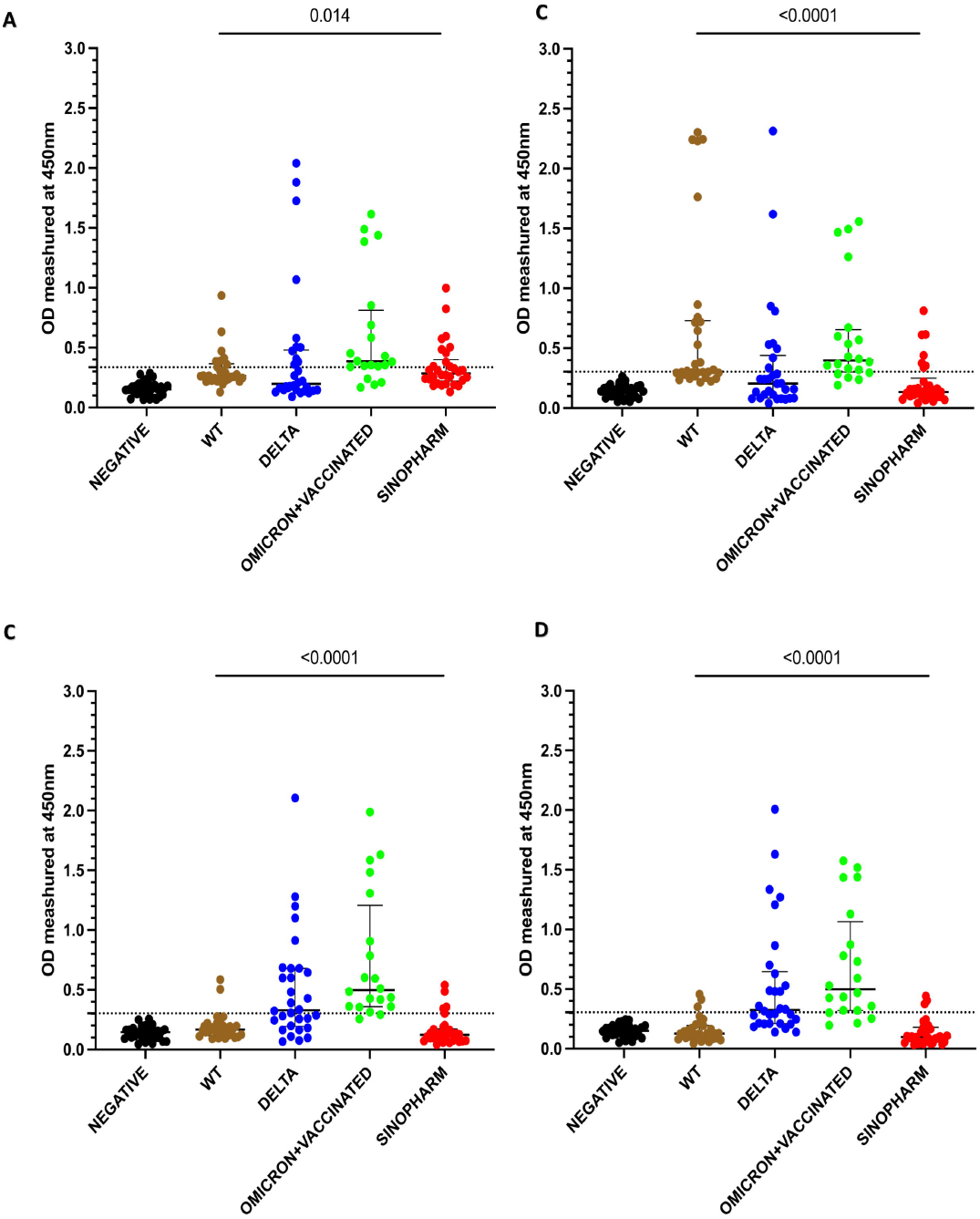
Antibody responses for the four overlapping pools of peptide of the N protein of SARS-CoV-2. IgG antibody responses were measured by an in-house ELISA for pool 1 (A), pool 2 (B), pool 3 (C) and pool 4 (D) containing overlapping peptides of the N protein in SARS-CoV-2 seronegative (non-vaccinated) individuals (n=30), WT infected individuals (n=30), delta infected individuals (n=30), omicron infected and vaccinated individuals (n=20), and Sinopharm vaccinees (n=30). Kruskal-Wallis test was used to determine the differences between the levels of antibody responses in four peptide pools (pool 1, 2, 3, and 4). Dotted line shows the cutoff value (OD value) of a positive response for the each of the peptide pools. The error bars indicate the median and the interquartile ranges. Black: seronegatives ; Brown: WT infected individuals ; Blue: Delta infected individuals ; Green: Omicron + vaccinated; Red: Sinopharm vaccinees.

Of those in who were infected prior to being vaccinated, the WT infected individuals had the highest positivity rates for pool 2, while delta infected individuals gave highest antibody responses to pool 3 and 4. Sinopharm vaccinees had the highest positivity rates for pool 1. However, Sinopharm vaccinees had overall lower positivity rates and magnitude of responses for all four peptide pools compared to naturally infected individuals.

### Mapping of antibody responses in the different cohorts to identify immunodominant regions of N protein

As the WT infected individuals (cohort B) had the highest responses to pool 2, Sinopharm vaccinees (cohort E) to pool 1, delta infected individuals (cohort C) to pool 3 and 4, we proceeded to map the immunodominant regions within these different pools of overlapping peptides, by testing antibody responses to these individual peptides separately.

In cohort D and E, the highest responses were observed for the two overlapping peptides 5 and 6 of pool 1 (peptides 1 to 15) (Supplementary Figure 1A and 1B). Cohort B (WT infected individuals) and D (omicron infected+ vaccinated) had the highest responses to overlapping peptides 23 and 24 of pool 2 (Peptide 16 to 30) (Supplementary Figure 1C and 1D). Individuals from cohort C (delta infected) and D, had the highest responses to overlapping peptide 40 and 41 of pool 3 (Peptide 31 to 45) (Supplementary Figure 1E 1F). In cohort C and D, the highest responses were observed for peptide 53 and peptide 54 of pool 4 (Peptide 46 to 59) (Supplementary Figure 1G and 1H). Based on these results, overlapping peptides 5 and 6 of pool 1, overlapping peptides 23 and 24 of pool 2, overlapping peptides 40 and 41 of pool 3 and overlapping peptides 53 and 54 of pool 4, were the immunodominant regions within the N protein.

### Characterizing antibody responses to the immunodominant regions identified within N protein

In order to further characterize the antibody responses to the above immunodominant regions within the N protein, the overlapping peptides 5 and 6, 23 and 24, 40 and 41 and 53 and 54 were pooled together from four different pools. The antibody responses for these regions were assessed in all cohort (cohort A to E), to identify responses in all individuals for these pools. Although the WT infected individuals gave the highest antibody responses to P23/24 (Figure 2A and B), there was no significant difference (p=0.057) in the magnitude of responses for the four immunodominant peptide pools in this group. Similarly, delta infected individuals had a similar magnitude of responses for the four pools (p=0.53). Omicron infected+vaccinated (cohort D) had the highest responses to P40/41 and P53/54, while Sinopharm vaccinees had the highest responses to P5/6 (Figure 2A and B).

**Figure 2:**
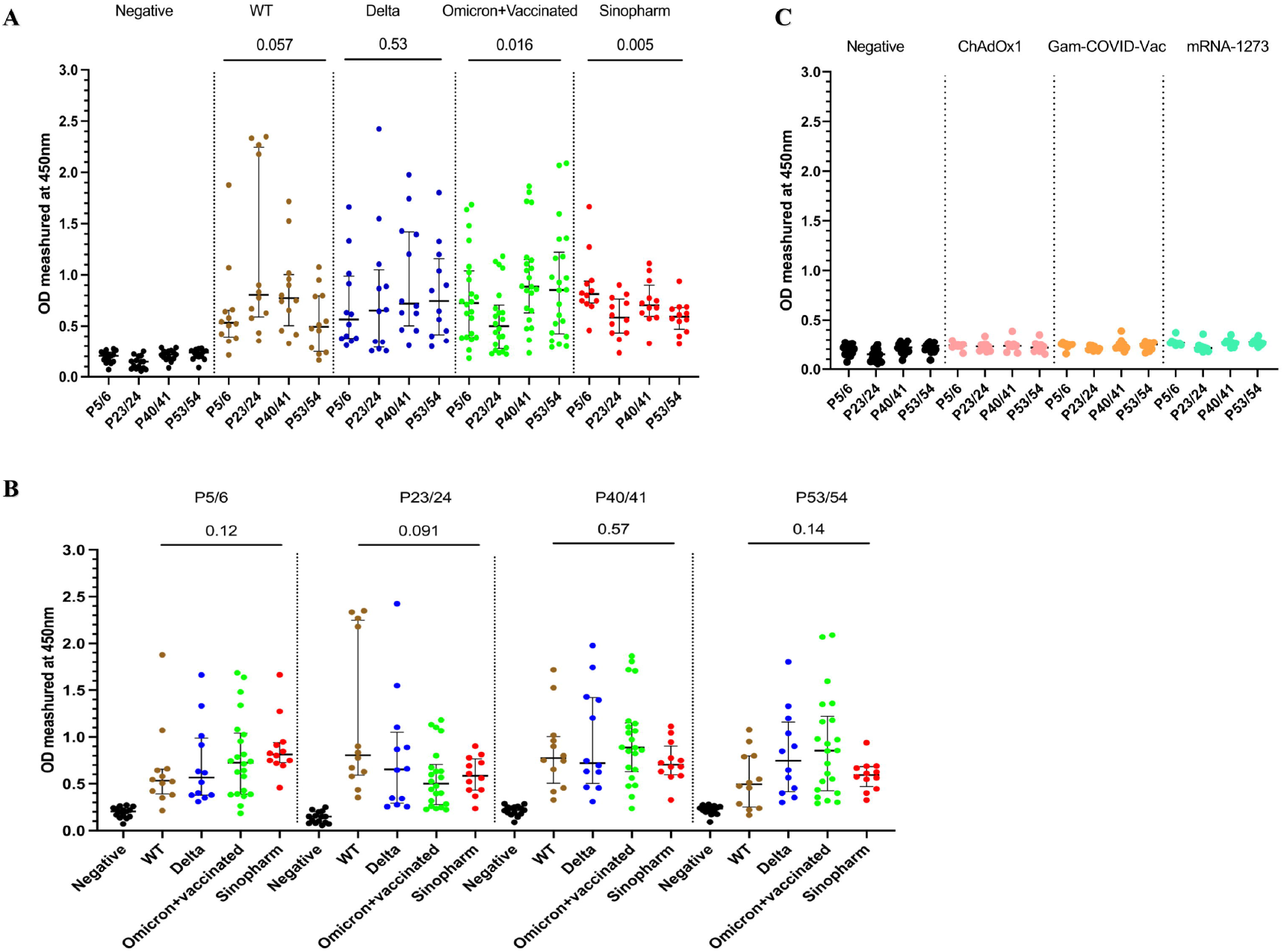
Characterizing antibody responses to the immunodominant regions identified within N protein. IgG antibody responses to the four immunodominant regions (P5/6, P23/24, P40/41, and P53/54) in the N protein were measured by an in-house ELISA in SARS-CoV-2 in SARS-CoV-2 seronegative (non-vaccinated) individuals (n=15), WT infected individuals (n=12), delta infected individuals (n=12), omicron infected and vaccinated individuals (n=22), and Sinopharm vaccinees (n=12) (A). The magnitude of antibody responses to these regions in the above cohorts were compared with each other (B). In order to determine specificity, antibody responses were measured in individuals who were vaccinated (uninfected) with AZD1222 (n=10), Moderna (n=10), and Sputnik V (n=10). Kruskal-Wallis test was used to determine the differences between the levels of antibody responses in four peptide pools (pool 1, 2, 3, and 4). The error bars indicate the median and the interquartile ranges.

The positivity rates for each of the four immunodominant regions in these cohorts is shown in table 2. Overall, all cohorts had >80% positivity rates for all 4 regions, except lower positivity rates in delta infected and omicron infected+ vaccinated individuals for P23/24 region. In contrast, all WT infected individuals gave a positive response for this region, while they had low positivity rates (66.7%) for P53/54.

**Table 2:** The number of individuals in different cohorts who gave a positive response to immunodominant regions within the N protein.

| SARS-CoV-2 Variant | Number of individuals who gave a positive antibody response |  |  |  |  |
| --- | --- | --- | --- | --- | --- |
|  | P5/6 | P23/24 | P40/41 | P53/54 | At least for one pair |
| WT (n=12) | 10 (83.3%) | 12 (100%) | 11 (91.7%) | 8 (66.7%) | 12/12 (100%) |
| Delta (n=12) | 11 (91.7%) | 9 (75%) | 11 (91.7%) | 10 (83.3%) | 12/12 (100%) |
| Omicron+ vaccinated (n=22) | 20 (90.9%) | 16 (72.7%) | 20 (90.9%) | 18 (81.8%) | 20/22 (90.9%) |
| Sinopharm (n=12) | 12 (100%) | 11 (91.7%) | 11 (91.7%) | 11 (91.7%) | 12/12(100%) |
| Total (n=58) | 53 (91.4%) | 48 (82.8%) | 53 (91.4%) | 47 (81.0%) | 56/58<br>(96.5%) |
| AstraZeneca (n=10) | 0 | 1 (10%) | 1 (10%) | 0 | 1/10 (10%) |
| Sputnik V (n=10) | 0 | 0 | 1 (10%) | 0 | 1/10 (10%) |
| Moderna (n=10) | 1 (10%) | 1 (10%) | 0 | 0 | 2/10 (20%) |
| Total (n=30) | 1 (3.3%) | 2 (6.7%) | 2 (6.7%) | 0 | 4/30 (13.3%) |

We also assessed specificity of the antibody responses to the four immunodominant regions of the N protein (P5/6, P23/24, P40/41, and P53/54), by investigating responses in those who received COVID-19 vaccines with only the spike protein. Accordingly, responses were assessed in those who received AZD1222 (ChAdOx1) (n=10), Moderna (mRNA-1273) (n=10), and Sputnik V (Gam-COVID-Vac) (n=10). None of the SARS-CoV-2 seronegative individuals (cohort A) or those who received ChAdOx1, Gam-COVID-Vac or mRNA-1273 responded to the peptides P53/54, while one individual (1/30) had a positive response to the peptides P5/6. Two individuals (2/30) responded to the peptides P23/24 and P40/41 (Table 2, Figure 2C). Therefore, while the specificity of peptide P53/54 was 100% in detecting SARS-CoV-2 N protein specific responses following or vaccination (whole virus vaccine) or infection, the specificity of peptides P5/6 was 96.7% while for peptides P23/24 and P40/41 it was 93.3%.

### Conservation of immunodominant regions of the N protein of SARS-CoV-2 with seasonal human coronavirus and SARS-CoV-2 variants of concern (VoC)

As the consensus peptide sequence may not be representative of the infecting subtype, we determined the conservation within these four immunodominant regions within the different SARS-CoV-2 variants (Alpha, QVX37034.1; Beta, QWW93444.1; Gamma, QXF23757.1; Delta, UKA47847.1; Omicron, (BA.1 (SriLanka/aicbu4450/2022), BA.2 (SriLanka/aicbu4463/2022) and BA.5 (USA/CA-CDPH-FS27225444/2022) and also the cross reactivity with other seasonal human coronaviruses (OC43, QBP84763.1; HKU1, ABG77571.1; NL63, YP_003771.1). We used Jalview software [28] and tools available at European Bioinformatics Institute (EBI) (www.ebi.sc.uk, 22 March 2022) to determine conservation between identified four immunodominant regions of the wild type SARS-CoV-2 (YP_009724397.2) The four regions in which the conservation and cross reactivity were assessed are as follows;

P5/P6: ^29^NGERSGARSKQRRPQGLPNNTASW^52^

P23/P24:^155^AAIVLQLPQGTTLPKGFYAEGSRG^178^

P40/P41:^274^FGRRGPEQTQGNFGDQELIRQGTD^297^)

P53/54: ^365^PTEPKKDKKKKADETQALPQRQKK^388^

All regions showed <50% sequence identity with the three seasonal human coronaviruses (Supplementary table 1 Figure 3A to 3D). P5/6 showed <20% sequence identity with OC43, HKU1, and NL63, with P53/54 showing <10% sequence identity with these viruses (Supplementary table 1). In contrast, the regions P23/24 and P40/41 showed >45% of sequence identity with OC43 and HKU1 but not with NL63. All four immunodominant regions were found to be conserved in both alpha and beta variants (Supplementary table 1 and Figure 3E to H), while there was 95.8% sequence identity with delta (single amino acid replacement) in the P53/54 region and 95.8% in the P40/41 region with the gamma variant (single amino acid replacement). All three omicron sub-lineages (BA.1, BA.2 and BA.5) P5/6 have a 3 amino acid deletion within the regions represented by P5/6 and therefore, a sequence identity of 87.5% (Figure 3E). P23/24 regions was 100% conserved in all five VoC (Figure 3F).

**Figure 3:**
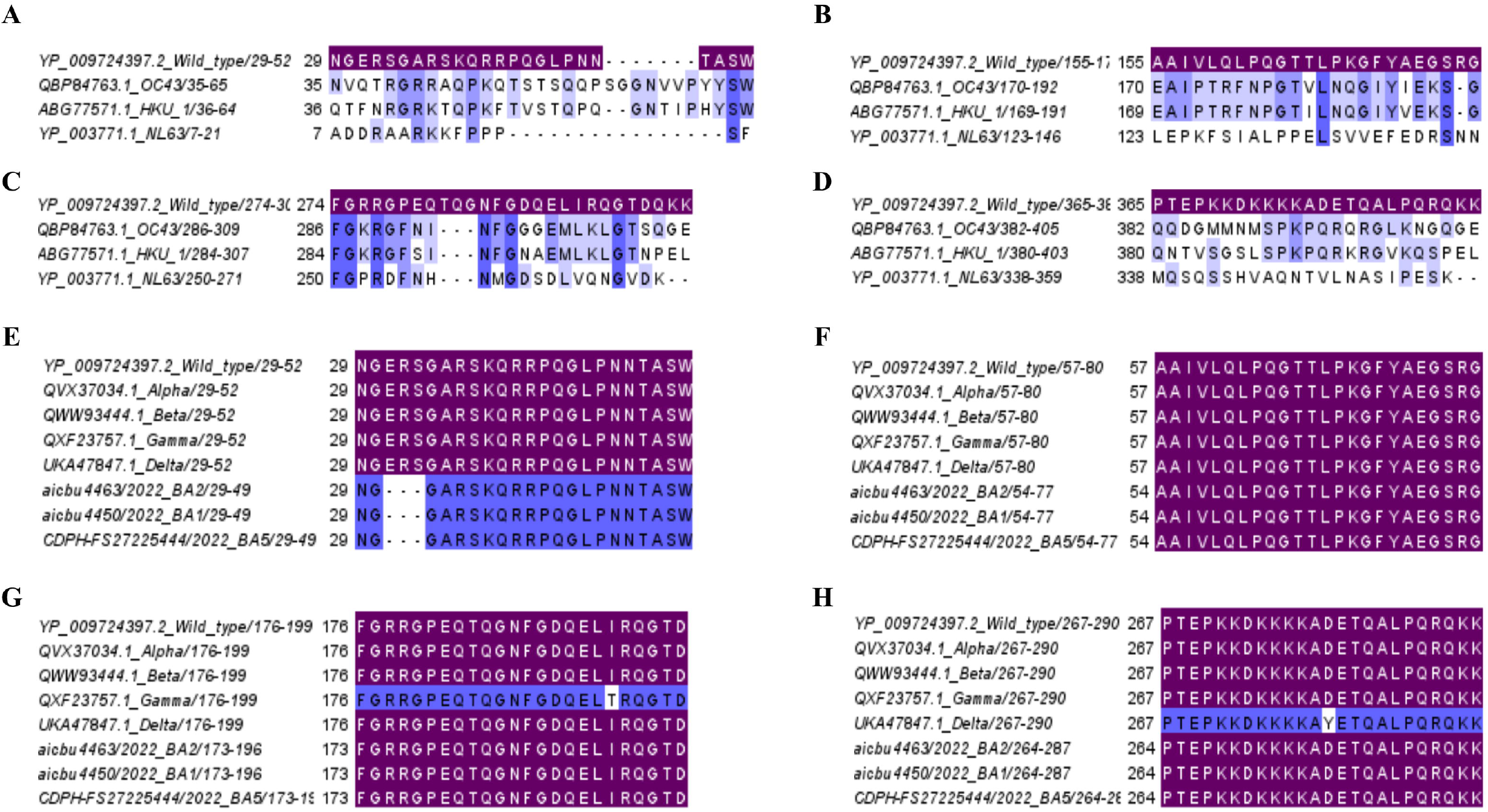
Analysis of conservation of immunodominant regions of the N protein of SARS-CoV-2 with seasonal human coronavirus and SARS-CoV-2 variants of concern (VoC) The cross reactivity of the four immunodominant regions P5/6 (A), P23/24 (B), P40/41 (C), and P53/54 (D) with three seasonal human corona viruses (OC43, HKU1, and NL63) were determined The conservation within these four immunodominant regions were also assessed for the five VoCs (alpha, beta, gamma, delta, and omicron (BA.1, BA.2, and BA.5). P5/6 (E), P23/24 (F), P40/41 (G), and P53/54 (H). Matching (sequence identity 100%) respective immunodominant regions were highlighted purple color.

As the SARS-CoV-2 virus continues to further evolve and due to the future threat of other bat coronaviruses spilling over and causing future pandemics, many Pan-Sarbecovirus vaccines are currently under development [9,19]. Therefore, we proceeded to find out the conservation of these four regions with 4 bat coronaviruses, RS4081, WIV1, RatG13 and Rf1 (Figure 4) and Supplementary table 1. These four regions showed >91% sequence identity with all the four bat coronaviruses.

**Figure 4:**
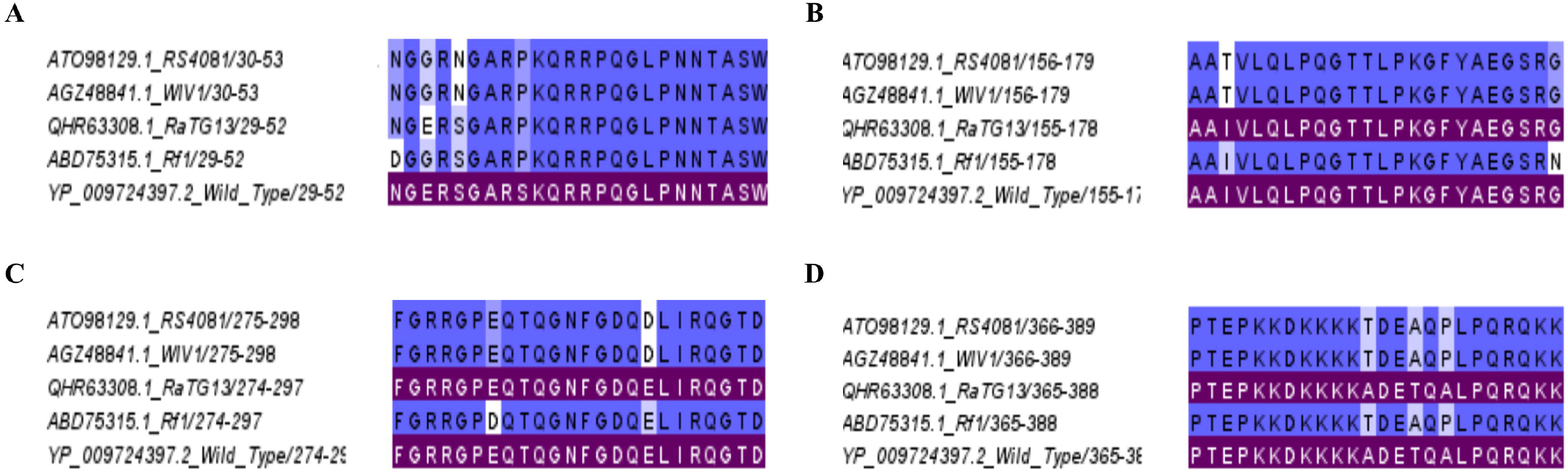
Analysis of conservation of immunodominant regions of the N protein of SARS-CoV-2 with bat coronaviruses. The cross reactivity of the four immunodominant regions P5/6 (A), P23/24 (B), P40/41 (C), and P53/54 (D) RS4081, WIV1, RatG13 and Rf1 were analyzed. Matching (sequence identity 100%) respective immunodominant regions were highlighted purple color.

### Antibody responses to the four immunodominant regions in patients with varying severity of COVID-19 due to the WT virus

We then sought to compare antibody responses between mild and severe disease during early and late stage of the infection in individuals who had mild illness (n=16) or severe illness (n=9) during acute stage (<7 days since onset of symptoms) and during late infection (21 to 28 days since onset of symptoms). Those with severe illness had significantly higher antibody responses to P23/24, P40/41 and P53/54 during the first week of illness compared to those with mild illness (Figure 5A). During late infection, those with severe disease had significantly higher antibody responses to all four regions than individuals with mild illness (Figure 5B).

**Figure 5:**
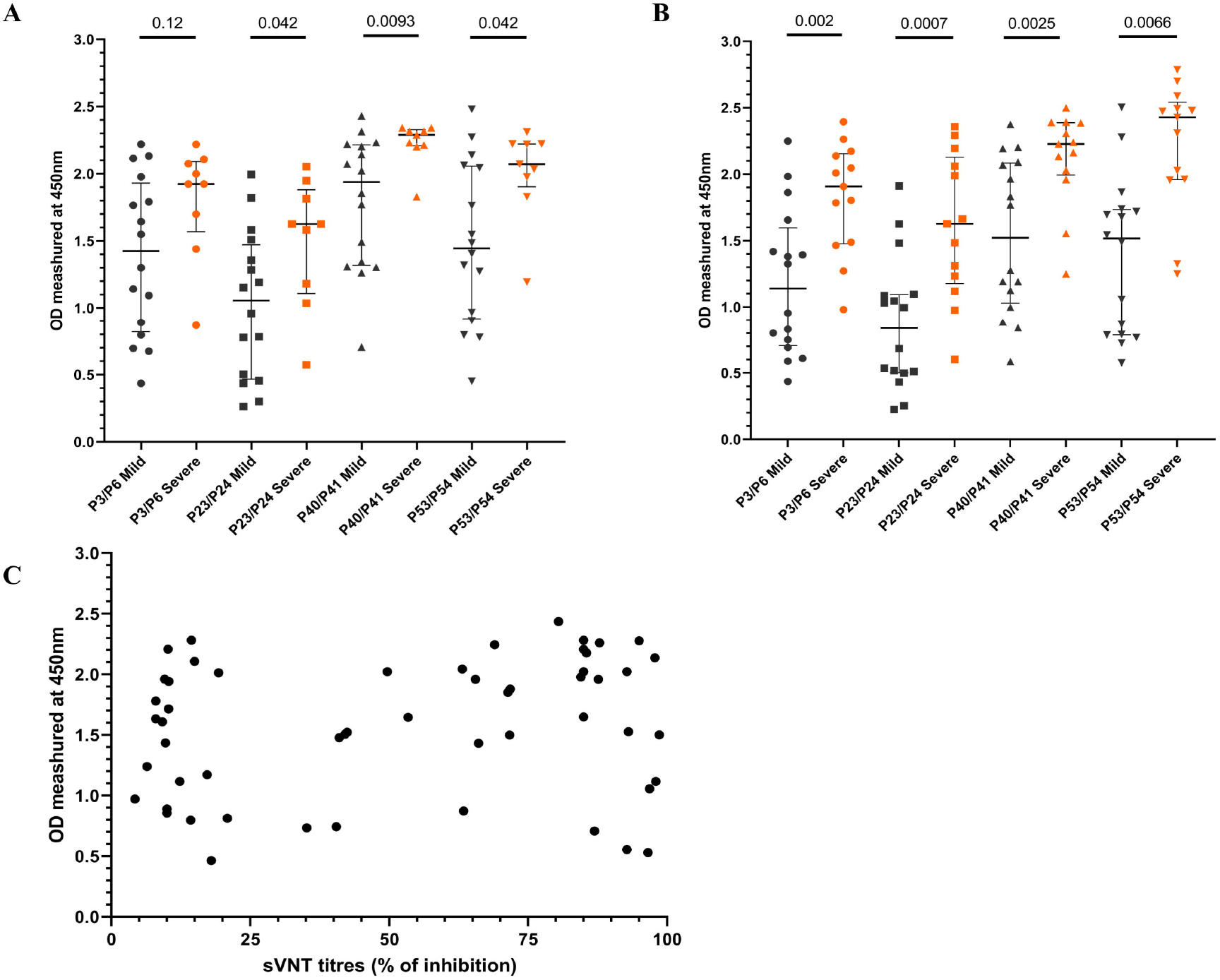
Antibody responses to the four immunodominant regions in patients with varying severity of COVID-19 due to the WT virus. Antibody responses to the four immunodominant regions were measured by an in-house ELISA, in patients infected with the WT of SARS-CoV-2 with mild (n=16) and severe disease (n=9) during early illness (<7 days since onset of symptoms) (A) and late illness (21 to 28 days since onset of symptoms (B). The antibody responses to the four regions were correlated with ACE2 blocking antibodies measured by the surrogate virus neutralization test, and the ACE2 blocking antibodies did not correlate with the levels of the four immunodominant regions (C). The Mann-Whitney U test (two-tailed) was used to determine the differences in antibody levels between those with mild and severe disease. All tests were two sided. The error bars indicate the median and the interquartile ranges.

We then sought to explore if antibody responses to these immunodominant regions, correlate with neutralizing antibody (Nab) responses, by comparing ACE2 blocking antibodies (which were shown to correlate with Nabs), by using a surrogate SARS-CoV-2-neutralizing antibody assay. The ACE2 blocking antibodies did not correlate with antibody response against all four regions (Spearman’s r=0.17, p=0.02) (Figure 5C).

## Discussion

In this study we have identified four immunodominant regions within the N protein, which gave a high frequency of responses in those infected with the WT virus, delta, omicron and those vaccinated with Sinopharm. Overall, >80% of individuals gave responses above the positive cut-off threshold to many of the four regions, with some differences with individuals who were infected with different VoCs. These regions were found to be 100% specific, as none of the seronegative individuals gave any responses. However, 6.7% to 10% of individuals who had received the spike proteins vaccines and therefore, should not have responses to these regions also responded. Although they had tested negative for infection with SARS-CoV-2 by the commercial N protein-specific antibody assay, it is possible that they could still have been naturally infected. Due to the high sensitivity and specificity of these regions, they have a potential to be used to identify N protein specific antibody responses, especially regions aa 29 to 52 (P5/6) and aa 365 to 388 (P40/41), for which >90% of individuals responded to. However, as we used overlapping peptides to identify potential epitopes, we are likely to have missed conformational epitopes which could be key antibody recognition sites.

The five domains of the N protein have shown to bind to RNA and carry out multiple functions including RNA interference, regulating virus replication and host immune evasion [6]. Of the four regions identified here, one is within the N terminal domain (aa 29-52, P5/6), one in the RNA binding domain (aa 155-178, P23/24), one within the dimerization domain (aa 274 to 297, P40/41) and one in the C-terminal domain (aa365 to 388/ P53/54). Some previous studies had identified immunodominant regions of the N protein, in llamas (domesticated South American camel), and had identified antibodies that bind to highly conserved regions within the N protein [32]. One of these antibodies had shown to bind to the C-terminal domain (aa 49-174) and two antibodies to the N-terminal domain (aa 247-364 and aa 365-419)[32]. The four immunodominant regions that were identified here, also fall within these three regions. Another study, which screened for B cell epitopes within the N protein using mouse models, identified aa 401 to 408 as the main antibody target. Our data show that, the predominant B cell epitopes identified within the N protein differs based on the infecting SARS-CoV-2 variant. For instance, those who were infected with the WT virus, predominantly recognized the aa155 to 178 regions (P23/24), whereas those who were infected with delta had the highest responses to the aa 29 to 52 and aa 274 to 294 regions. However, the responses to these overlapping peptides were assessed using the sequence of a Wuhan virus strain isolated from USA in 2020 and the epitope recognition could be different, based on the sequence of peptides used. In those who were infected with different omicron sub-lineages (BA.1 and BA.2) also had the highest responses to the aa 29 to 52 and aa 274 to 294 regions. Therefore, the predominant B cell epitope recognition, appears to differ based on the variant of infection. Although Sinopharm vaccinees responded to all four regions, the magnitude of the responses was significantly lower than following natural infection. This is possibly due to natural infection inducing more robust responses to the N protein than following inactivated vaccines containing the whole protein.

The mortality rates and hospitalization rates have varied widely throughout the COVID-19 pandemic in different countries, with many countries in Europe, and United States reporting higher mortality rates and hospitalization rates than some countries in Africa and Asia, despite higher rates of vaccination [13,14]. These differences could be attributed to reporting of COVID-19 deaths and limitations in testing, as many countries in sub-Saharan Africa have reported high excess mortality rates [10]. Sri Lanka experienced high mortality rates during the delta outbreak during the months from June to October 2021 prior to vaccination [24]. However, mortality rates have been significantly less (0.85/ million individuals in Sri Lanka) during the massive omicron wave (BA.2), than many European countries and the United States (mortality rates 4.03/million individuals in Europe and 5.4/million individuals in United States) [13]. Only 18% of Sri Lankans had received an mRNA booster dose, when the omicron variant was rapidly spreading in Sri Lanka [14]. The lower mortality rates seen in Sri Lanka during the omicron outbreak were unlikely to be due to under reporting or limited testing as the excess mortality rates in Sri Lanka were found to be less than the excess mortality rates reported in Europe and North America [10]. Sinopharm/BBIBP-CorV was the most widely used vaccine, in Sri Lanka with 12 million (70.6%) individuals receiving this vaccine by end of December 2021 [9]. Sinopharm/BBIBP-CorV vaccine was found to be less immunogenic than the mRNA-1273, AZD1222 and Sputnik V, 3 months post second dose, in a head-to-head comparison in the Sri Lankan population, based on ACE2 blocking antibodies and antibodies to the receptor binding domain of the spike protein [15]. However, as Sinopharm/BBIBP-CorV is an inactivated vaccine, it did induce T cell and antibody responses to the N protein [16]. In addition, although the N protein was thought to be localized to the cytosol, it was recently shown that this protein was expressed on the surface of infected cells [20]. As the N protein has shown to bind to several different types of chemokines, antibodies against the N protein could also inhibit chemotaxis of leucocytes [20]. Furthermore, antibodies bound to N protein were shown to activate FcR expressing innate immune cells, further contributing to the phagocytosis and apoptosis of infected cells[20]. Although there could be many reasons for the differences in mortality rates for different variants, it is possible that antibody responses to the N protein, offered additional protection in Sinopharm vaccinees, which should be further investigated.

Due to the rapidly evolving nature of SARS-CoV-2 and emergence of more immune evasive omicron sub-lineages, there is a global effort to develop a pan-Sarbecovirus vaccine [7,9,11]. However, many pan-Sarbecovirus vaccines only use the spike protein as the immunogen and explore the immune responses to the spike protein [19,25], while only a few vaccines also include the N protein [9]. In this study we show that the four immunodominant regions identified here, were highly conserved regions within SARS-CoV-2 and the bat coronaviruses. Although the neutralizing antibody responses for many bat Sarbecoviruses has been investigated [27], there are no data if antibodies targeting the main B cell epitopes within the N protein, also cross-neutralize the most frequent bat Sarbecoviruses, which would be important. While vaccination, especially with an mRNA booster dose induced high levels of Nabs and protected individuals from severe disease [4], natural infection and vaccination induced a high magnitude of durable immune responses [2]. In fact, it was shown that two doses of an mRNA vaccine and natural infection gave similar immune responses as three doses of a mRNA vaccine, while the immune responses induced by natural infection were longer lasting [2]. Therefore, in order to induce persistent and broad immune responses, antibody and T cell responses to the N protein may play an important role in addition to Nabs.

## Conclusions

We have identified four immunodominant regions within the N protein of SARS-CoV-2, which are highly conserved in the SARS-CoV-2 variants and also show high conservation in bat Sarbecoviruses. Responses to these regions were highly specific and elicited responses in > 90% of naturally infected individuals or those who received a whole virus inactivated vaccine to at least two of the regions. As these regions were highly specific with high sensitivity, they have a potential to be used to develop diagnostic assays and to be used in development of vaccines.

## Data Availability

All data produced in the present work are contained in the manuscript

## List of abbreviations

Nabs: Neutralization antibodies
N protein: Nucleocapsid protein
VoC: variants of concern
sVNT: surrogate virus neutralizing test
RBD: receptor binding domain
WT: Wuhan strain of SARS-CoV-2

## Ethics approval and consent to participate

Ethics approval was obtained by the Ethics Review Committee of the University of Sri Jayewardenepura. All individuals gave informed, written consent.

## Consent for publication

Not applicable

## Availability of data and materials

All data is available in the manuscript and figures.

## Authors’ contributions

Conceptualization of the study: PDP, CJ, GNM

Data curation: ISA, TN, JJ, TR, HK, SD

Project administration: CJ, AW, GNM

Experiments: PDP, FB, DM, LP

Data analysis: PDP

Funding: CJ, GSO, GNM

Writing the manuscript: PDP, GNM

Reviewing the manuscript: CJ, GSO

## Acknowledgements

None

## Supplementary figure legends

**Supplementary Figure 1**: **Mapping of antibody responses in the different cohorts to identify immunodominant regions within the pools of overlapping peptides**.

IgG antibody responses were measured by an in-house ELISA for individual overlapping peptides of pool 1 (peptide 1 to 15) in omicron + vaccinated (A) and Sinopharm vaccinees (B), pool 2 (peptide 16 to 30) in WT infected (C), and omicron+ vaccinated (D), pool 3 (peptide 31 to 45) delta infected (E) and omicron + vaccinated (F) and in pool 4 in delta infected(G) and omicron+ vaccinated (H). 10 individuals were included in each cohort to identify individual antibody responses. The error bars indicate the median and the interquartile ranges.

