## Supplementary figures and images for "Identification of differences in the magnitude and specificity of SARS-CoV-2 nucleocapsid antibody responses in naturally infected and vaccinated individuals"

### supplementary figures

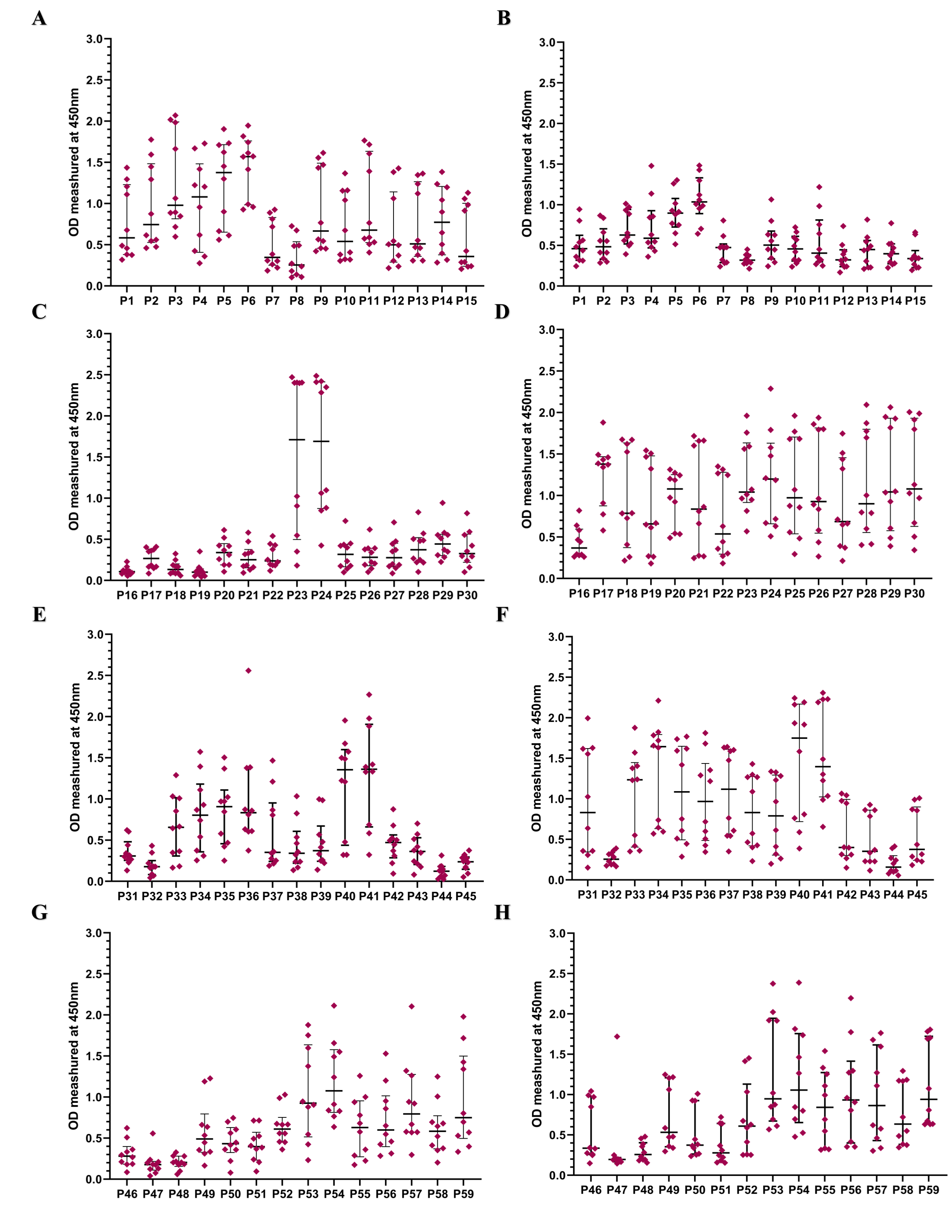
