## Supplementary tables for "Identification of differences in the magnitude and specificity of SARS-CoV-2 nucleocapsid antibody responses in naturally infected and vaccinated individuals"

| Human Corona viruses | Percentage of sequence identity (%) | | | |
| --- | --- | --- | --- | --- |
|  | P5/6 | P23/24 | P40/41 | P53/54 |
| OC43 | 16.7 | 45.8 | 45.8 | 8.3 |
| HKU1 | 12.5 | 45.8 | 45.8 | 8.3 |
| NL63 | 12.5 | 8.3 | 25.0 | 4.2 |
| VoCs | P5/6 | P23/24 | P40/41 | P53/54 |
| Alpha | 100 | 100 | 100 | 100 |
| Beta | 100 | 100 | 100 | 100 |
| Gamma | 100 | 100 | 95.8 | 100 |
| Delta | 100 | 100 | 100 | 95.8 |
| Omicron  (BA.1, BA.2, and BA.5) | 87.5 | 100 | 100 | 100 |
| RS4081 | 95.8 | 95.8 | 95.8 | 91.7 |
| WIV1 | 95.8 | 95.8 | 95.8 | 91.7 |
| RatG13 | 95.8 | 100.0 | 100.0 | 100.0 |
| Rf1 | 95.8 | 95.8 | 95.8 | 95.8 |

**Supplementary table 1: Conservational analysis of the four immunodominant regions of the N protein with human corona viruses and SARS-CoV-2 VoCs**
